# Instantaneous Three-Dimensional Scanning for Foot Orthosis Design: Clinical Validation of a Multicamera Photogrammetry 3D Scanner

**DOI:** 10.64898/2026.05.13.26352176

**Authors:** Joshua A Taylor, Alexander J Terrill, Aaron Wholohan, Renee Nightingale, Ollie Nagle, Edmund Pickering, David W Holmes, Sean K Powell, Maria A Woodruff

**Author notes:** Authors Joshua Taylor and Alexander J Terrill contributed equally to the work and are joint first authors.

## Abstract

3D scanners have revolutionised how podiatrists capture foot morphology in order to design custom orthoses (insoles). While various 3D scanning technologies are used in clinical practice, they vary greatly in cost and ease of use and many of these are not specifically designed for podiatry applications. There is limited literature comparing accuracy between scanners, and many approaches require prolonged scan times during which the patient must remain still. Multicamera photogrammetry offers a promising solution by enabling high-quality, rapid 3D scanning which other devices cannot provide.

This study compared the accuracy and clinical utility of four 3D scanners. One was a high accuracy reference scanner (Artec Spider) which was used as a gold standard. Two further scanners which are commonly used in the clinic were also investigated (Apple iPad 6 with Structure Sensor attachment ‘iPad’, and Envisic VeriScan Podiatric Scanner ‘laser’) and these were directly compared with a novel prototype multicamera photogrammetry 3D scanner. The left feet of 20 healthy volunteers were scanned using each of the four devices and scans were evaluated for accuracy, completeness, and acquisition and processing times.

All scanners produced clinically acceptable scans, with the novel photogrammetry scanner demonstrating superior accuracy. Scan times varied significantly between scanners, with the photogrammetry device capturing scans much faster. All scanners had acceptable levels of completeness, though the iPad and photogrammetry outperformed the laser scanner.

These results provide a valuable tool for clinics seeking guidance on scanner selection and highlight the benefits of instantaneous photogrammetry scanning to improve workflow efficiency and accessibility.

## 3 Introduction

Foot pain is experienced by approximately 13–36 % of the general population (1) and has been linked with an increased risk of falls (2), impaired mood (3), and reduced quality of life (4). Populations that include runners (5), the elderly (6), and those with comorbidities, such as osteoarthritis (7) and rheumatoid arthritis (8), experience an increased risk of foot pain, highlighting the diverse nature of people who may seek pain management. Custom foot orthoses (CFOs) that are produced based upon the individual’s foot morphology and function are one treatment modality practitioners use to assist people with foot pain (6,9), meaning the accurate and timely capture of human foot morphology is highly important in clinical practice.

To manufacture CFOs, foot-health practitioners (including podiatrists) would traditionally obtain negative casts of patient foot morphology using plaster bandage or foam impression boxes. These processes can be time consuming, messy, or potentially error-prone (10). These casts would be filled with plaster to produce a positive cast, which would be modified to achieve the desired foot orthosis shape. Finally, a thermoplastic would be vacuum formed to the modified positive cast and finished to produce the orthosis. With the advent of computer aided design and computer aided manufacture, this process has become digitised. A model of the foot is now commonly obtained using 3D scanning, computer aided design tools are used to create the orthosis shape, and 3D printing or computer numerically controlled (CNC) milling is used to fabricate the orthosis. Compared to orthoses produced based on a foam impression cast, orthoses manufactured from a direct scan are associated with improved adherence, satisfaction, as well as lower costs (11).

Three-dimensional scanning of the foot is the crucial first step in this digitised process. 3D scanners commonly used clinically employ structured light, laser time-of-flight, or laser triangulation techniques (12). Structured light scanners project a light pattern onto a surface, and the reflected light is recorded by a camera which is offset from the projector, and triangulation is used to obtain depth information (13). Laser time-of-flight scanners project infrared light onto a surface and depth information is determined how long it takes for the signal to return (13). Laser triangulation scanners project a laser and measure the angle of reflection to determine depth information (13). A common challenge for these three methods of 3D scanning is that they require the light source or sensor to be moved around the foot to capture the surface from different perspectives, which can take several minutes, and should the foot move, the scan fails. Consequently, the foot must be held stationary throughout the scan, and any movement would necessitate the scan to be restarted. If the 3D scan could be taken instantaneously (as fast as a photograph) then the person moving would not pose a challenge and the process would be faster, more accurate and easier for both the patient and the clinician and would save significant time and resources.

Despite the widespread use of 3D scanning in manufacturing foot orthoses, there has been limited research comparing different types of 3D scanners for this purpose. Within the existing research, there is a large diversity of 3D scanning hardware, software, protocols, and approaches to statistical analysis (12). A systematic review by Farhan et al (10) in 2021 compared 3D scanning to traditional methods of capturing foot and ankle morphology prior to CFO manufacture. The review found 3D scanning was faster than casting methods – especially for experienced users – with comparable accuracy and reliability in the context of CFO fabrication (10). Farhan et al. (14) evaluated the speed and accuracy of seven 3D scanners in capturing foot, ankle and lower limb morphology prior to ankle-foot orthosis manufacture and identified significant differences in accuracy of clinical measures and speed of scanning, with three scanners not meeting a minimum threshold for accuracy with disparities exceeding 5%. Rogati et al (15) compared a high resolution laser scanner to a low-cost structured light scanner (based on a Microsoft Kinect sensor) in weight bearing positions, and determined accuracy was suitable for clinical applications. Nickerson et al. (16) compared flatbed laser triangulation and hand-held structured light scanners, and determined the impressions obtained from laser scanners – which were used in a partial weight bearing configuration – had flatter and less pronounced arches and greater compression of the soft tissues than those produced by the hand-held scanner (which was used in a non-weightbearing configuration). Similarly, Chhikara et al. (17) evaluated a structured light scanner, flat-bed laser triangulation scanner, and a smartphone-based laser time-of-flight scanner in both non-weight bearing and partial weightbearing positions and reported minimal difference in the models produced between scanners, with the positioning of the foot having a greater impact on orthosis design.

An alternative to existing scanners is photogrammetry, which was first reported to be used to create a 3D model of a foot in 1981 (18). Photogrammetry uses multiple photographs captured from different perspectives to obtain depth information (19). The photographs can be obtained by a single camera, with images sequentially taken from different perspectives (single camera photogrammetry), or by multiple cameras positioned around an object that are triggered to capture images simultaneously (multicamera photogrammetry) (20). A key opportunity provided by multicamera photogrammetry is that simultaneous image capture can enable near-instantaneous 3D scanning. While single-camera photogrammetry has been demonstrated as capable of constructing models of the foot (14,21,22), there has been no research identified that compares multicamera photogrammetry to alternative 3D scanning methods for foot capture.

Therefore, the purpose of this study was to compare two 3D scanners that are commonly used in clinical practice and a novel multicamera photogrammetry-based 3D scanner against a reference high-accuracy structured light scanner. Key outcomes of interest are accuracy, completeness, scanning acquisition time, and scan processing time.

## 4 Materials and Methods

### 4.1. Study design

This study followed a repeated-measures validation study design. To streamline inter-study comparisons, where possible, this follows the Consistent Reporting Three-dimensional scanning (CRITIC) checklist proposed by Allan et al. (12). Institutional ethics approval was obtained from QUT’s University Human Research Ethics Committee (ethics approval number 6316).

### 4.2. Participants

This study included 20 healthy adult participants. Sample size was calculated using statistical software G*Power (23) (Heinrich Heine University Düsseldorf, Germany). Using accuracy results (1.5 mm ± 1.1 mm) from a study by Nightingale et al. (24) that benchmarked multi-photogrammetry scanning of the face against the Artec scanner, a minimum sample size of 10 participants was required to generate a study power of 95%, however, high interest during recruitment allowed the participant cohort to reach n=20.

Participant recruitment took place from the Queensland University of Technology (QUT) podiatry community. Participants were eligible if they were 18 years or older, were able to lie in the supine position for 45 minutes, and the foot and ankle had adequate range of motion to allow positioning for scanning. Participants were excluded if they were non-ambulatory, had active foot or lower-limb ulceration, amputation, or movement disorders affecting the ability to remain stationary.

### 4.3. 3D scanning devices

Four scanning devices with software unique to each device (Table 2) were used to capture foot data, including a high-resolution scanner, two scanners that are commonly used by podiatrists, and a novel prototype photogrammetry scanner.

**Table 1:** Scanning devices. Summary of scanning devices (3D scanning principle, scan position, and software).

| **3D scanner** | **Artec Space Spider (Version 1)** | **iPad 6 with Structure Sensor Mk1** | **Envisic Veriscan laser scanner** | **Photogrammetry device** |
| --- | --- | --- | --- | --- |
| **3D scanning principle** | Structured light | Structured light | Red-line laser triangulation | Multicamera Photogrammetry |
| **Operation** | Handheld | Handheld | Mounted flatbed | Handheld |
| **Processing software** | Artec Studio 13 (Artec Group, Luxembourg) | 3DsizeME (TechMed 3D Inc., Canada) | LaserCAM Prescribe (LaserCAM Orthotics, Adelaide, Australia) | Meshroom (AliceVision, France) |

**Table 2:** Summary of Accuracy and Completeness results for all scanners. Results are reported as mean ± standard deviation. p value for RMS error calculated using repeated measures ANOVA; p value for Completeness calculated using repeated measures ANOVA of logit transformed data.

|  | Artec -<br>Photogrammetry | Artec -<br>Smoothed<br>Photogrammetry | Artec - Ipad | Artec - Laser |  |
| --- | --- | --- | --- | --- | --- |
| n | 19 | 19 | 19 | 19 | p |
| **RMS error (mm)** |  |  |  |  |  |
| **Plantar Foot** | 0.25 ± 0.06 | 0.26 ± 0.07 | 0.34 ± 0.06 | 0.63 ± 0.13 | <0.001 |
| **Rearfoot** | 0.14 ± 0.04 | 0.14 ± 0.05 | 0.29 ± 0.08 | 0.70 ± 0.24 | <0.001 |
| **Midfoot** | 0.25 ± 0.06 | 0.26 ± 0.07 | 0.29 ± 0.07 | 0.47 ± 0.07 | <0.001 |
| **Completeness (%)** |  |  |  |  |  |
| **Plantar Foot** | 98.9% ± 1.4% | 98.6% ± 1.5% | 98.4% ± 1.1% | 96.6% ± 1.3% | <0.001 |
| **Rearfoot** | 99.5% ± 0.6% | 99.3% ± 0.9% | 98.9% ± 1.0% | 97.1% ± 1.8% | <0.001 |
| **Midfoot** | 98.4% ± 2.1% | 97.9% ± 2.4% | 97.9% ± 1.7% | 96.4% ± 1.9% | <0.001 |

**Table 3:** Acquisition and processing times. Mean and standard deviation displayed for acquisition time and processing time (seconds) for 3D scans (n=20).

|  | Acquisition Time (s) |  |  |  | Processing Time (s) |  |  |  |
| --- | --- | --- | --- | --- | --- | --- | --- | --- |
|  | mean | SD | min | max | mean | SD | min | max |
| **Photogrammetry** | <1 |  |  |  | 414 | 49 | 352 | 554 |
| **iPad** | 60 | 12 | 35 | 81 | 29 | 5 | 20 | 38 |
| **Laser** | 24 | 1 | 23 | 26 | 0 |  |  |  |
| **Artec** | 301 | 80 | 153 | 445 | 600 | 179 | 332 | 940 |

The Artec Space Spider 3D Scanner (Figure 1a) is a gold high-resolution metrology rated structured light 3D scanner (‘Artec’, Artec Group, Luxembourg). The Artec was used as a control as it has been widely used in literature as a control scanner for comparison studies (24–28) and has a reported 3D point accuracy of 0.05 mm (29).

**Figure 1:**
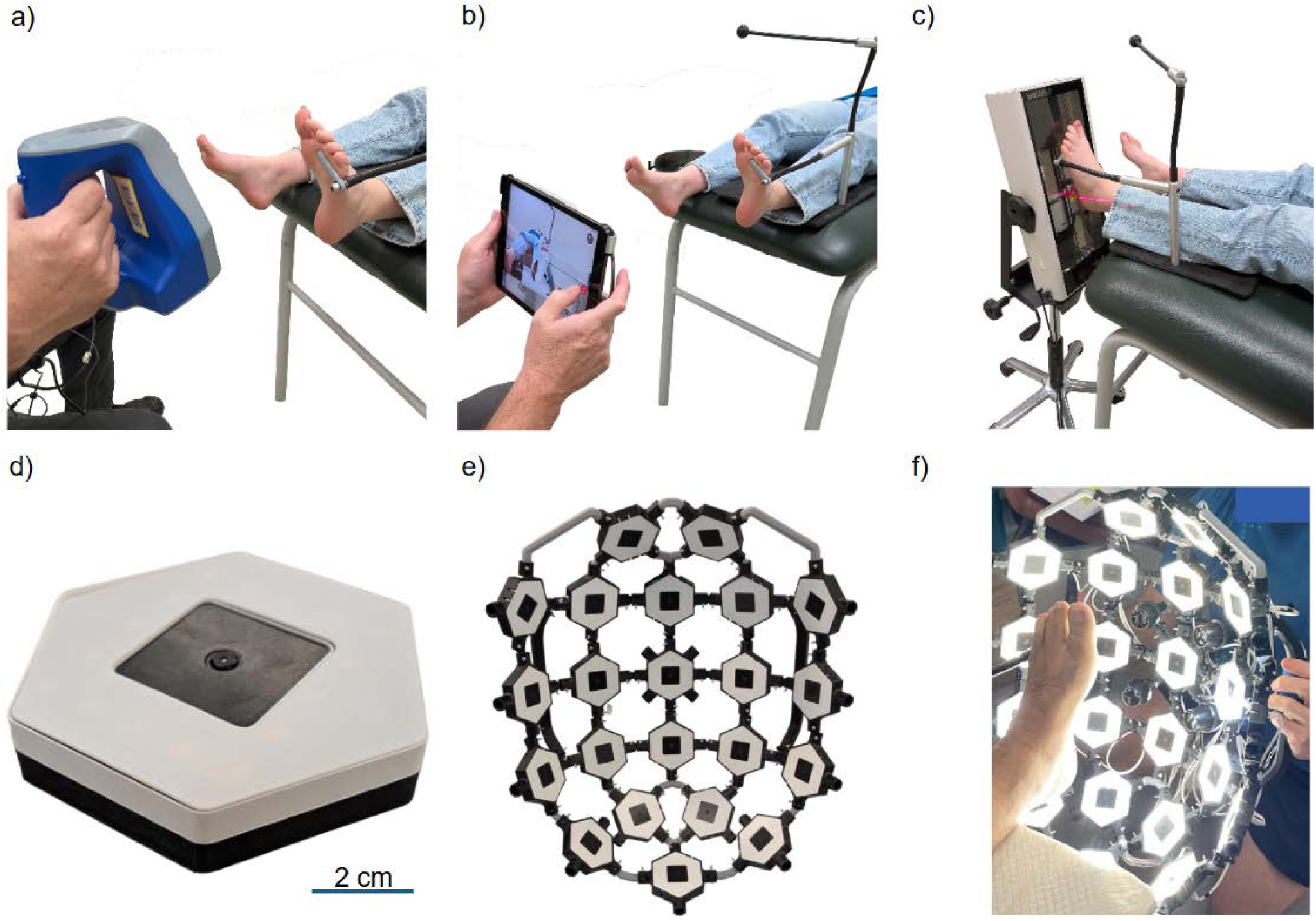
3D scanners used in study and positioning when scan data is acquired. (a) Artec Spider in use; (b) iPad 6 with structure sensor attachment in use; (c) Envisic Veriscan in use; (d) Photogrammetry module; (e) Photogrammetry rig purposively designed for 3D scanning the plantar foot, incorporating 22 scanning modules; (f) Photogrammetry scanner in use

Two scanners were selected that are commonly used in clinical practice by podiatrists in Australia. First, an Apple iPad 6 (Apple Inc., Cupertino, CA, USA) with Structure Sensor 1 attachment (Structure, Boulder, Colorado, USA) (Figure 1b). Second, an Envisic VeriScan Podiatric Scanner (‘laser’, Envisic LLC, Albuquerque, NM, USA) (Figure 1c).

The final scanner was a multi-camera photogrammetry scanner (‘photogrammetry’, QUT, Brisbane, Australia). The photogrammetry scanner was a prototype research device developed by the research team, comprising 22 camera modules spatially distributed around the foot that could be triggered to capture images synchronously (Figure 1d). Each module consisted of a Raspberry Pi Zero and a PiCamera 2 NOIR, as well as an array of LED lights providing 250 Lumens of light. Camera modules were held in a 3D printed frame that was designed to ensure coverage of the plantar surface of the foot, medial and lateral borders, and posterior heel (Figure 1e-f). The scanning frame included converging targeting lights that would indicate correct positioning of the scanner around the foot.

### 4.4. 3D scanning protocol

The bare left foot of each participant was scanned with the participant in a supine-lying position on a clinical plinth, using all four scanning devices. The Artec was used first as the control scan, then the order of the test scanners used was randomised for each participant (30). Unilateral scans were performed to limit participant and investigator fatigue. Prior to scanning, talcum powder was applied evenly to the foot of each participant to reduce specular reflection and improve tracking when using the Artec. No markers were placed on the skin (12). Each participant was scanned with the photogrammetry rig three times (with the first scan used for analysis) and the iPad and laser scanner were used once.

A foot stabilising device (ScanBud, KLM Laboratories Inc., Valencia, California, USA) was used to stabilise the foot during scanning. The foot was supported by the stabilising device, with the supporting bar positioned plantar to the 2^nd^ - 5^th^ metatarsophalangeal joints, the subtalar joint in neutral position, and midtarsal joint pronated, as shown in Figure 2. To minimise movement, participant feedback was gained prior to the scanning procedure regarding comfort level and perceived stability on the device with minor positional adjustments of the foot made accordingly. The stabilising device, as delivered, uses a transparent supporting bar not compatible with the Artec 3D scanner. As such, this was replaced with an opaque 3D printed bar. Following placement of the foot on the stabilising device, participants remained in this position for the duration of all scanning. To assess for participant movement, five participants were rescanned using the Artec following the completion of all experimental scans.

**Figure 2:**
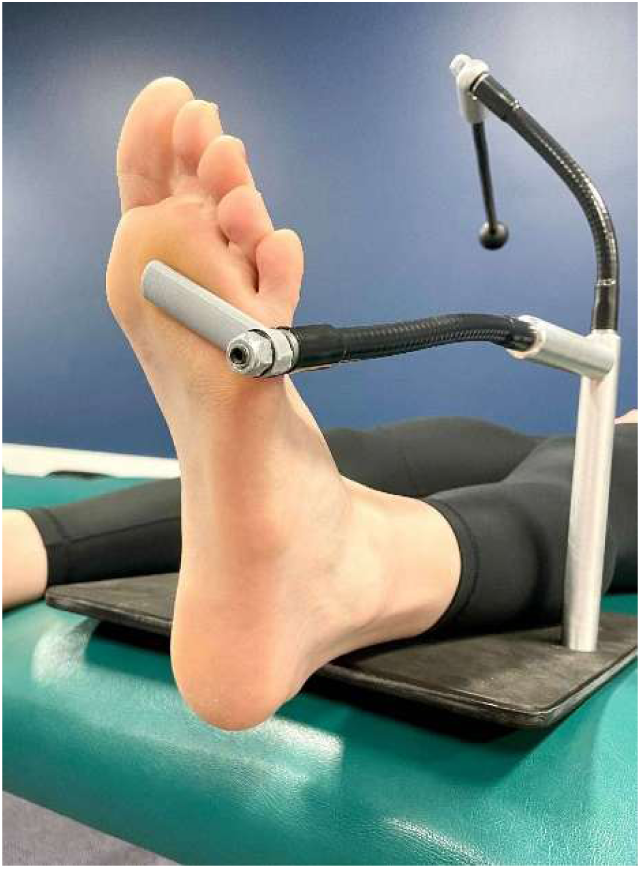
Foot stabilising device and participant positioning. A Foot stabilising device with bare left foot of a representative participant resting in scan position and heel overlying end of plinth

The foot of each participant was scanned to capture the plantar surface, medial arch, lateral border, and posterior heel. The dorsal surface of the foot and toes were not considered relevant for this study as they are unimportant for custom foot orthosis manufacture; therefore, time was not spent ensuring their accurate capture. In addition to the scanned foot, portions of the foot stabiliser were unavoidably captured.

Due to their different scanning approaches each scanning device was operated and data processed according to a standard operating procedure:

1. **Artec Spider scanner**: in a seated position, the investigator moved the handheld scanner around the foot at a distance of 0.2–0.3 m (Figure 1a). Visual feedback on scan progression was available on a nearby Dell Precision 7760 laptop (Intel Core i7-11850H CPU, Nvidia GeForce RTX 3080 Laptop GPU, 32 GB RAM). The recorded data was processed in Artec Studio 13 (Artec Group, Luxembourg) using the autopilot tool and saved as a stereolithography (STL) file.
2. **iPad with Structure Sensor**: in a seated position, the investigator moved the iPad around the foot at a distance of 0.5–0.6 m (Figure 1b). Visual feedback on scan progression was available on the iPad screen through the iOS application 3DsizeME (TechMed 3D Inc., Canada). The recorded data was transferred to MSoft Windows software (TechMed 3D Inc., Canada), aligned, excess data was cropped, and the model was reconstructed and saved as a .STL file.
3. **Laser scanner:** housed in a supportive bracket, the laser scanner was positioned parallel to the plantar surface of the foot at a distance of 0.01–0.02 m (Figure 1c). Data acquisition was performed using LaserCAM Prescribe software (LaserCAM Orthotics, Adelaide, Australia) and visual feedback on scan the outcome was available on a nearby Microsoft Surface Laptop 3 (Intel Core i5-1035G7 CPU, Intel Iris Plus GPU, 8 GB RAM). Scans were exported as WRL files which were then converted to STL files using MeshLab (31) (version 2022.02., Visual Computing Lab, Italy).
4. **Photogrammetry rig scanner:** the investigator held the scanner aimed at the plantar surface of the foot with the visual targeting system used to aid in rig positioning (Figure 1f). When targeted, the investigator used the trigger to activate 3D scanning, which simultaneously captured images from each of the cameras. Images were transferred to a high performance Dell Precision 5820 computer (Intel Xeon W-2295 CPU, Nvidia RTX A5000 GPU, 128 GB RAM) on which the images were processed to create a 3D model using Meshroom (version 2023.2.0, AliceVision, France), with feature extraction using the DSP-SIFT and AKAZE algorithms, the ULTRA setting for describer quality and density, and no depth map downscaling. 3D images were exported from Meshroom as an OBJ file, and imported to MeshLab for STL file conversion.

### 4.5. Segmentation

Following data acquisition, models were imported into CloudCompare (version 2.12.4) and segmented using the ‘segment’ tool to isolate anatomically relevant regions, resulting in three separate models: the plantar surface, the rearfoot and the midfoot (shown in Figure 3). As segmentation is a manual process, all segmentation was performed by a single researcher for consistency. The plantar foot model represents the parts of the foot that are most relevant to orthosis design, and excludes the toes and stabilising device. For the plantar foot model, the imported model was cropped approximately 5 mm distal to 1st metatarsophalangeal joint on the medial side, and proximal to stabilising device on the lateral side. The lateral border and rearfoot were cropped to 28 mm depth from the most plantar point. The highest point of medial longitudinal arch cropped to 40 mm, tapering to 28 mm at both the distal and proximal boundaries of the arch. From this plantar foot model, separate rearfoot and midfoot models were derived to allow quantification of accuracy in different regions of the foot. The rearfoot comprised the posterior 30% of the total foot length, and the midfoot comprised the remaining portion of the plantar model.

**Figure 3:**
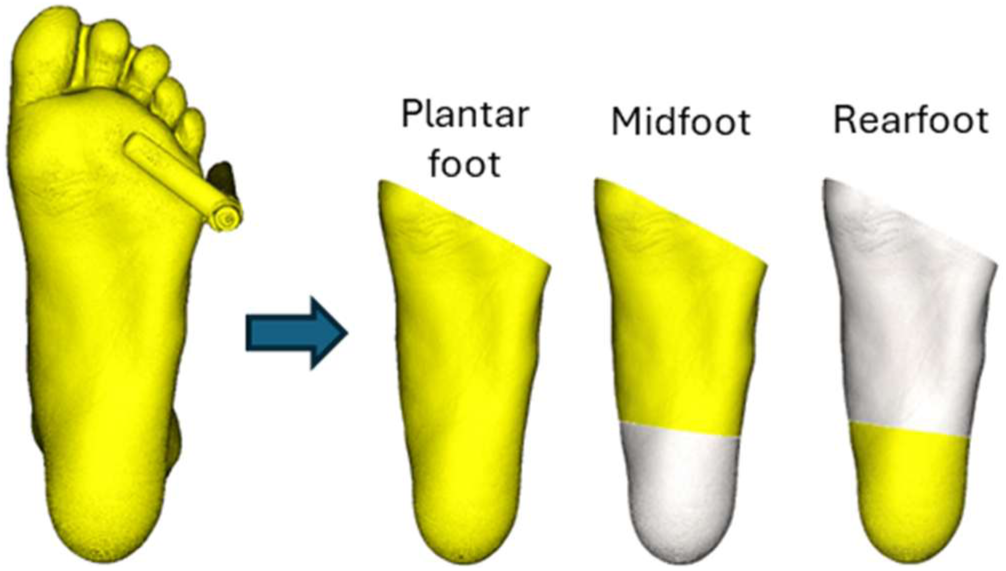
Segmentation of the 3D scans. Each scan was segmented into the plantar foot, and sub-segmented to the rearfoot and midfoot portions of the plantar foot.

After segmentation, photogrammetry models were smoothed using MeshLab Taubin smooth function (32). This smoothing was performed due to surface noise visible on the unsmoothed models.

### 4.6. Accuracy

For this study *accuracy* is defined as the Root Mean Square (RMS) error in millimetres (mm) of 3D points between the experimental scans and the control scan, whereby a larger reported error indicates reduced accuracy. Accuracy was calculated in accordance with the established procedure outlined in Nightingale (24). Using CloudComPy (33) (a python wrapper for CloudCompare software), all experimental models were aligned to the comparable reference scan using iterative closest point (ICP) registration, and the cloud-to-mesh tool used to calculate the deviation between each 3D point in the aligned segmented scans and the closest 3D point in the reference Artec scan. For each comparison, the root mean square of all deviation values were calculated in Python. Visualisation Toolkit (.VTK) files were exported to Paraview (34) (version 6.0.0, Kitware Inc., Clifton Park, NY, USA) to create colour maps representing the deviation between the control and experimental scans.

### 4.7. Completeness

*Completeness* was defined in this study as the percentage of vertices of the experimental scans that were within 2 mm of the corresponding closest vertex in the control scan. A result of 100% would indicate all vertices of the Artec scan were ‘captured’ in the experimental scan. Completeness was calculated using the ‘cloud-to-mesh’ feature in CloudComPy, with the Artec set as the comparison scan and the scan being analysed set as the reference scan (24). 3D vertices in the Artec scan that were within 2 mm of the experimental scan were considered ‘captured’ (24).

### 4.8. Acquisition Time

This study defined *acquisition time* as the amount of time (in seconds) needed for each scanner to complete data acquisition once activated by the investigator. This is the time, during which a patient in a clinical setting would need to remain stationary. It does not include the time required to position the participant or position the scanner prior to scanning commences. If repeat scans were required due to a user error, the acquisition time was restarted. Acquisition times were measured with a stopwatch at time of acquisition.

### 4.9. Processing Time

*Processing time* is defined as the amount of time (in seconds) required to generate a 3D model file from the corresponding acquired scan file. This does not include the time taken to transfer data between computers, or the time taken to convert a mesh file of a different format to an STL file usable by CloudCompare for analysis. Processing times were measured with a stopwatch at the time of processing.

For the Artec, processing time was the time required to process the data using the autopilot pipeline. For the iPad, processing time was the time required to load the scan data in mSoft, align, crop and reconstruct the model. For the Photogrammetry rig, processing time was the time to run the photogrammetry pipeline in Meshroom. No processing time was required for the laser scanner, as a .WRL mesh file was exported immediately after the acquisition was completed.

### 4.10. Statistical analysis

SPSS statistics V30 (IBM Corporation Armonk, NY, USA) was used for statistical analysis.

The accuracy and completeness data for one participant was excluded as an outlier. For this participant, scans obtained using the second and third scanners (laser and iPad respectively) showed substantial deviation from the rest of the sample. This suggested significant foot movement between scans for this participant.

Accuracy and completeness data was tested for assumption of normality with the Shapiro-Wilk test. Normality of scanner accuracy data was observed following removal of the outlier. For completeness data, Assumption of normality of raw data was not met, hence data was logit transformed. Repeated measures ANOVAs were conducted. Mauchly’s test showed that the assumption of sphericity was not met for scanner accuracy or completeness, therefore Greenhouse-Geisser correction was used. Post-hoc pairwise comparisons t-tests were performed, with Bonferroni correction applied to control for multiple comparisons.

Statistical tests for completeness were confirmed by non-parametric analysis using the Friedman test and Post-hoc pairwise comparison was performed using Wilcoxon signed-rank tests with Bonferroni correction. P values <0.05 were considered statistically significant.

## 5 Results

Twenty participants were recruited for this study with a mean age of 32.4 ± 14.7 years. Ten participants were male, and ten were female. Complete data were collected for all participants, with one participant removed from analysis of completeness and accuracy as an outlier. When presented, results are mean ± SD.

### 5.1. Participant movement – Repeated Artec Scans

The repeated Artec scans produced average accuracies of 0.15 ± 0.06 mm, 0.06 ± 0.02 mm and 0.15 ± 0.06 mm for the plantar foot, rearfoot and midfoot, respectively. All Artec-Artec comparisons produced 100 % completeness. This indicates that a foot was held sufficiently stationary by the stabilising device during the scan process.

### 5.2. 3D Scanner Accuracy

Figure 4 and Table 2 show there was no significant difference between the accuracy of the smoothed and non-smoothed photogrammetry derived 3D scans (plantar foot p=0.20; rearfoot p=1.0; midfoot p=0.466). The smoothed photogrammetry scans were significantly more accurate than the iPad for the plantar foot (p=0.006) and rearfoot (p<0.001) segments, and had similar accuracy to the iPad in the midfoot segment (p=0.451). The smoothed photogrammetry scans were significantly more accurate than the laser scanner for plantar foot (p<0.001), midfoot (p<0.001), and rearfoot (p<0.001) segments. While the photogrammetry scanner was more accurate, the average accuracy of all scanners was within 2 mm. Colour maps (Figure 4) demonstrate the directional deviation between the experimental and control scans for one representative participant.

**Figure 4:**
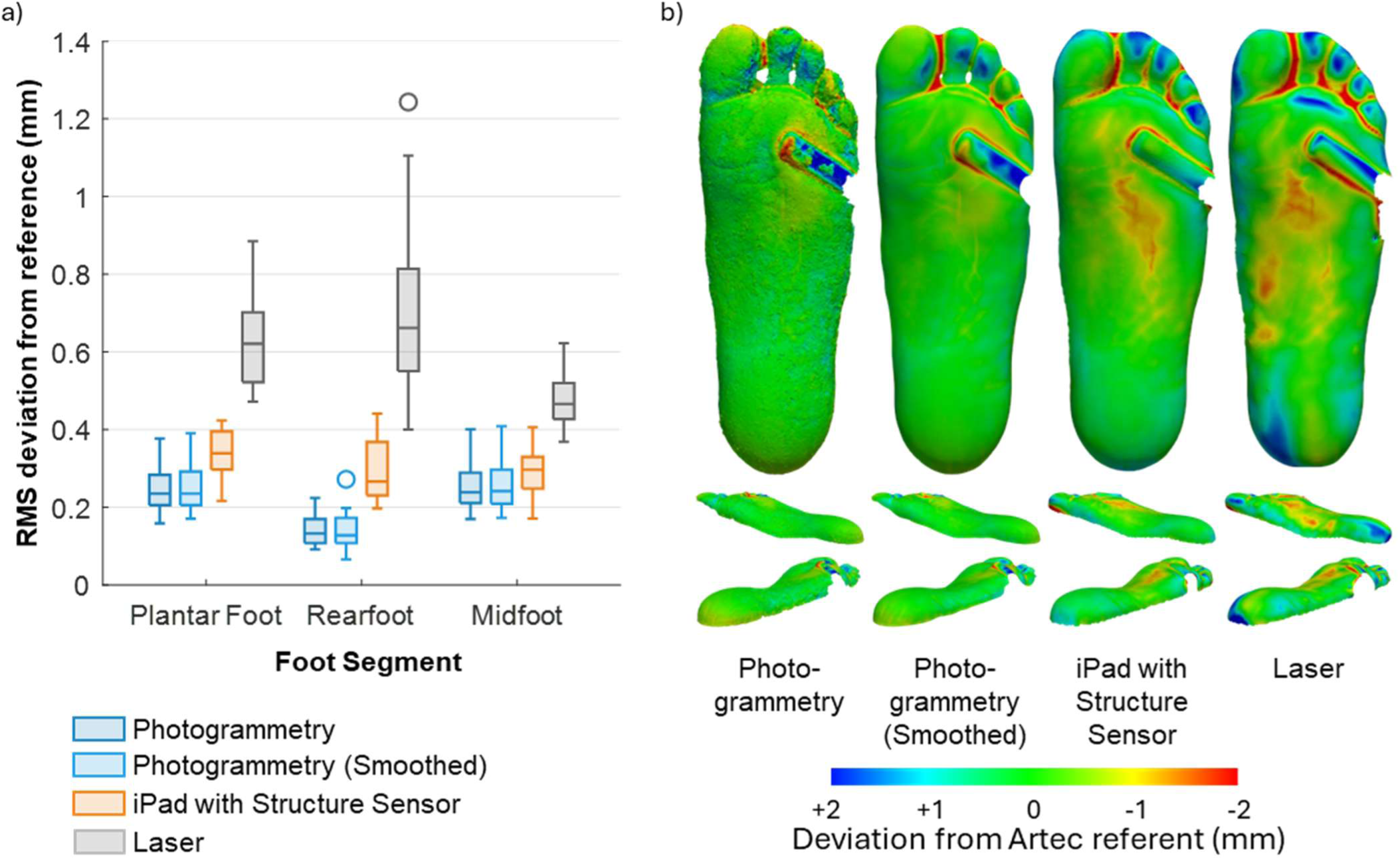
(a)Accuracy measured ass RMS deviation (mm) from Artec reference scanner (n=19). (b) RMS deviation distribution generated for one representative participant’s non-segmented foot. Colour represents deviation between experimental scan and reference Artec scan.

**Figure 5.**
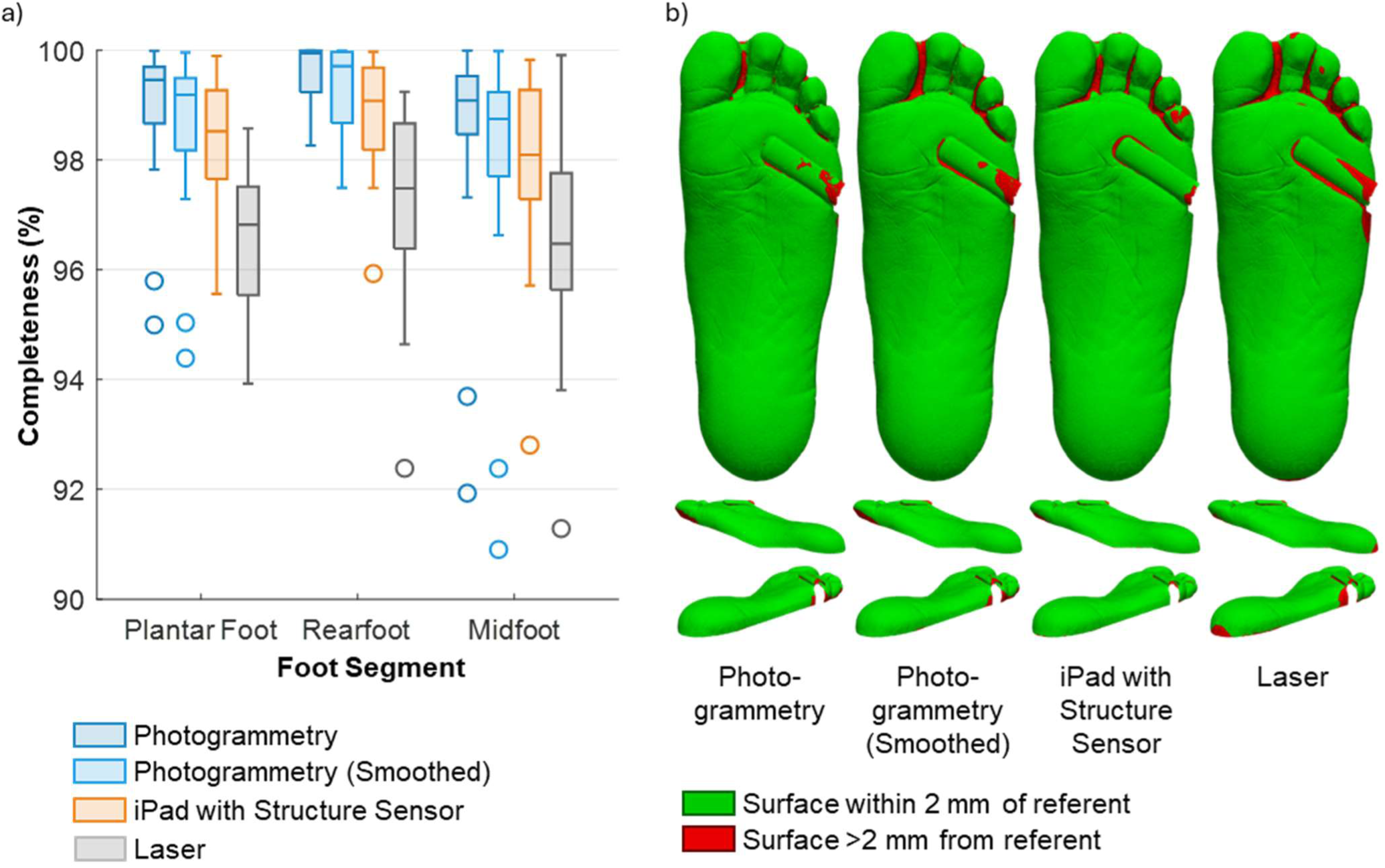
(a): Completeness of scans (%) compared to Artec Reference scanner (b) Completeness distribution generated for one representative participant foot. Colour representation of calculated distances between the nearest matching vertices in the experimental scans and the control for one representative foot prior to segmentation.

### 5.3. Completeness

Completeness was generally good across all scanners (Table 2). The smoothed photogrammetry scan had statistically significant lower completeness than the unsmoothed photogrammetry across all the plantar foot (p<0.001), rearfoot (p=0.007), and midfoot (p<0.001), however the magnitude of the mean difference was small. The iPad scanner had lower completeness than the photogrammetry scanner at the plantar foot: (p=0.04) and rearfoot (p=0.008), though the difference at midfoot did not reach statistical significance (p=0.22). The differences between the smoothed photogrammetry scans and iPad were not statistically significant at any sites (plantar foot: p=0.52; rearfoot: p=0.11; midfoot p=1.0). The Laser scanner demonstrated reduced completion compared to the photogrammetry scanner at all segments (plantar foot: p<0.001; rearfoot: p<0.001; midfoot p=0.012), the smooth photogrammetry scans at the plantar foot and rearfoot (plantar foot: p<0.001; rearfoot: p<0.001; midfoot p=0.083), and the iPad at plantar foot and rearfoot (plantar foot: p=0.004; rearfoot: p=0.006; midfoot p=0.11). On examination of completeness distribution, incompleteness was generally localised to the medial and lateral borders of the foot and posterior heel. When analysed using non-parametric statistical methods, the difference in completeness between the Laser and iPad at the midfoot also achieved statistical significance (p=0.01).

### 5.4. Acquisition and Processing Time

Acquisition and processing times (Table 4) were recorded for each of the four scanning devices. The photogrammetry rig always took less than 1 second to acquire the scan data but took between 352-554 seconds to create a model. The laser scanner always took 23-26 seconds to complete the scan and generate a model. iPad and Artec scanners demonstrated greater variance in acquisition time, with the iPad taking between 35-81 seconds to acquire the scan and 20-38 seconds to create the model, and the Artec taking 153-445 seconds to acquire the scan and 332-940 seconds to create the model.

## 6 Discussion

This paper has three key findings that advance the field of orthosis design using 3D scanning technology. First, we demonstrate that 3D scans with high levels of accuracy can be obtained using the iPad, laser triangulation, or photogrammetry approaches. Next, while all scanning methods can produce scans of sufficient accuracy for orthosis design, the novel prototype photogrammetry scanner achieved greater accuracy across all areas of the foot. Third, this enhanced accuracy was achieved though instantaneous scanning, a notable advantage over other methods that require longer capture times and risk patient movement abrogating the scans.

In this paper, while photogrammetry demonstrated greater **accuracy**, the RMS surface to surface deviation of all scanners was deemed sufficient for the design of foot orthoses. This outcome is relevant for clinical practice, as clinical 3D scanners have been widely adopted, and are already acknowledged as faster and capable of reducing material waste and costs compared to traditional plaster casting methods to capture foot morphology (10), and improving patient outcomes (11).

While all scanners showed acceptable accuracy overall, the laser scanner had a comparatively reduced accuracy around the rearfoot. Lower accuracy in this region observed with the laser scanner is likely due to the shallow angle between the surface of the skin and laser, which will reduce the depth resolution of the scanner. Conversely, greater accuracy of the photogrammetry 3D scanner in this region is likely due to the geometry of the purpose designed scanner. The scanner was designed with cameras intentionally positioned to maximise coverage of this part of the foot, deemed important for orthosis design by the clinicians in the design and research team. Likewise, good accuracy is observed with the iPad and Artec as both can be moved around the heel to capture it from different perspectives.

The finding that photogrammetry produced high accuracy scans is in contrast to Farhan et al. (14), which identified that models constructed using the smartphone-based single camera photogrammetry app Trnio had lower accuracy than other clinical 3D scanners. The improved result in the current study may owe to the specific design of the multicamera photogrammetry rig which utilised fixed camera positioning to maximise coverage of the foot without relying on the operator to ensure sufficient coverage, and further, allowed uniform lighting. The findings in the current study are supported by studies that have demonstrated good accuracy using photogrammetry for 3D scanning other areas of the body. Using handheld photogrammetry, Nightingale et al. (24) found photogrammetry accurate with RMS differences of 1.4 ± 0.5 mm in capturing facial morphology of 20 healthy participants, and in a later study (25), an RMS difference of 1.4 ± 0.3 mm compared to the Artec in capturing the external ear. In this study, we have found improved accuracy in capture of the foot, which could be because the foot is a less complex structure than the face and ear, which aids in accurate pairwise photo matching. These differences may also highlight the significant influence that the photogrammetry software pipeline has on the quality of the resultant model.

While accuracy is considered to be important in 3D scanning for foot orthoses, there is currently no clear clinical standard of a minimum accuracy threshold. Based upon the 3D scan, computer-aided design tools are used to create a model of an orthosis, which is then manufactured. The algorithms used by software to design a contoured orthosis from a 3D scan using these tools – as well as the inaccuracies introduced - are often not disclosed. Further, there are manual design steps, which lead to inter-designer differences that often exceed 1 mm (35). Given the inaccuracies inherent in the design process, sub-millimetric accuracy in 3D scanning is likely unnecessary, and all scanners compared in this study could produce a model with sufficient accuracy for foot orthosis design. This is in agreement with Chhikara et al (17), who indicated that scanner accuracy is less important than positioning the foot.

**Completeness** is a measure of whether all parts of a foot were captured with the experimental scanner. In this study, all devices demonstrated high levels of completeness, suggesting sufficient required data was present to then enable subsequent orthoses design and manufacture. Results in this study – ranging between 96.4–99.0% completeness for the plantar foot – exceed those found in Nightingale et al. (24) who reported 85 ± 15 % for facial scanning and Nightingale et al. (25) who reported 80 ± 11 % when scanning the external ear. This improvement is likely because the surface of the foot is less complex. In contrast, the ear and face contains complex surfaces that could obstruct the scanners’ line of sight.

In this study, the **acquisition time** of the photogrammetry rig was nearly instantaneous. This is in comparison to the iPad and Laser clinical scanners that took an average of 60 and 24 seconds respectively. The acquisition times recorded in this study parallel those recorded in previous studies where D’Ettorre et al. (36) found a multi-camera photogrammetry system captured facial data in 0.0015 ± 0.0 seconds. Furthermore, iPad acquisition time in the present study (60 ± 11 seconds) mirrored that of Powers et al. (37), where 60–90 seconds were needed to scan the lower limb for AFO fabrication, but was longer than described by Farhan et al. (14) where an average of 21 seconds were required and Farhan et al. (38) which required an estimated 30 seconds. The capture time taken for the Laser scanner is longer than the 7 seconds that is stated by the manufacturer (39) – This may be because the scanner was operated through third party software, which could define process parameters that prolong capture time.

In addition to taking consultation time, acquisition time is of considerable importance in clinical practice as a scan requires the patient must remain still, with any movement potentially causing a capture error and requiring the scan to be restarted. While we identified negligible movement occurred in this study of healthy adults, people with hyperkinetic movement disorders and children may not be able to remain still for the required capture duration. The near-instantaneous acquisition offered by photogrammetry addresses this concern and offers significant advantages over other scanners.

While acquisition time was much faster using multi-camera photogrammetry, the **processing time** was longer than other experimental scanners – despite being processed on a high-performance computer. Photogrammetry processing time is highly variable in literature, with fluctuations dependent on factors such as number of photographs taken (24), overlap between photograph pairs (40) and computer performance capability (25). Demonstrating this, Nightingale et al. (24) required 70 ± 15 minutes to process 40 photographs of the face, and in two external ear scanning studies, Ross et al. (20) required 24.6 ± 12 minutes to process 30 photographs and Nightingale et al. (24) required 41 ± 21 minutes..

The longer processing time demonstrated by photogrammetry is relevant, as the podiatrist must be able to visualise the 3D model to ensure adequate scan quality prior to discharging the patient. While the processing time is ‘hands-off’ meaning that it can occur in parallel to other clinical activities, given the average consultation in Australian podiatric practice is between 21–30 minutes (41), it is important that processing time does not cause delays. It is expected that optimisation of hardware and software could markedly improve processing time, and modern cloud-based processing infrastructure could enable access to high performance computing. The photogrammetry prototype hardware included many cameras – which likely contributed to a high accuracy result – but would also have extended the processing time. In a paper exploring the number of photos required to reconstruct a 3D scan of an ear, Ross et al. (20) demonstrated that increasing the number of photos from 30 to 90 resulted in a 9-fold increase in the processing time, therefore, optimising the number of cameras could lead to reduction in processing time. Furthermore, a significant time-component of the processing pipeline was the ‘structure from motion’ task, in which, the relative positioning of the cameras in space are determined through feature matching. This task could be eliminated if a more rigid frame were used with known relative camera positions. Finally, there is significant scope for optimisation of processing parameters within the photogrammetry pipeline that have not been explored in this study, such as feature extraction and mapping algorithms.

Several important study design features were considered to strengthen the reported outcomes. Firstly, this study undertook a sophisticated approach to measuring accuracy at a vertices-level analysis rather than using measures with less descriptive power, such as foot length and width. Next, minimal foot movement was noted across the participant cohort using the foot stabilising device, meaning differences in scanner accuracy and completeness can be primarily attributed to the capabilities of the devices. Third, the recruitment of asymptomatic participants lowers the barrier for future study replicability and provides a baseline for further research. Fourth, the randomisation of device order for each participant reduced the risk of investigator fatigue or participant movement affecting the results of one device disproportionately compared to other devices. Finally, in design and conduct of this study, we endeavoured to follow the CRITIC checklist developed Allan et al. (12), increasing the ability to streamline future inter-study comparisons.

Despite the results of this study being strengthened by several design features, limitations were present. While a strength from the perspective of providing a baseline for further research, restricting the study cohort to asymptomatic adult participants limits the generalisability of the results to sub-sections of the community commonly seen in podiatric practice. It would be valuable to purposively assess scanner utility in symptomatic and paediatric populations, as well as populations with differing skin tones and foot deformity.

Secondly, in this study, participant’s feet were captured in a non-weight bearing position using a commercial foot stabilising device. However, in clinical practice, there is no standardised approach to foot positioning. Partial-weight bearing and full weight bearing positions are frequently used and shown to produce significant differences in foot morphology and foot orthoses produced from the 3D scan (17,42). A limitation of the iPad, Artec, and photogrammetry rig in their current configurations is that they cannot be used to 3D scan in a partial weight bearing or weight bearing position. Further, many podiatrists will obtain scans in a non-weightbearing position without stabilising the foot. The high accuracy reported in this paper is in the context of the foot being stabilised – a non-stabilised foot is likely to lead to greater inaccuracy, particularly when using a scanner with a longer acquisition time.

Finally, an iPad 6 device and Structure Sensor Mark 1 was selected as one of the study scanners due to lack of access to newer models. It remains unclear how the latest devices would change the recorded results – while accuracy or speed improvements might be expected, in a study exploring scanning for ankle-foot orthoses, Farhan et al. (14) determined a Structure Sensor Mark 1 was 40% faster than the Structure Sensor Mark II with equivalent accuracy. Despite this, it is unlikely that clinicians would remain up to date with each hardware release, meaning these results still hold value in clinical practice.

While this study investigated the 3D capture of foot morphology in the context of fabricating orthoses and reducing foot pain, the need for accurate and timely foot acquisition is significantly broader. For example, contemporary 3D scanners may also have applications to optimise footwear fit (43), monitor wound healing (44), and could be used to facilitate enhanced care via telehealth for individuals in rural and remote clinical community settings. As scanning device capabilities improve and capital expenditure requirements reduce, the scope of scanning devices will only expand.

Multicamera photogrammetry offers exciting opportunities to advance clinical practice. Instantaneous 3D scanning eliminates the need for patients to remain still, which may be especially beneficial when scanning the feet of children and individuals with movement disorders. In addition to capturing foot morphology instantaneously, multicamera photogrammetry can simplify the 3D scanning process. The prototype design integrated targeting lights that guided optimal scanner positioning, consistently producing high-quality scans for every participant. Unlike hand-held structured light scanners, which require a skilled operator, the photogrammetry rig demands minimal expertise, making it a more accessible tool. This ease of use could enable wider access to 3D scanning for foot orthoses - for example, in the context of a podiatry workforce shortage in Australia, this could allow allied health assistants or other health care workers in remote areas to collaborate with podiatrists via telehealth to provide foot orthoses to patients in underserved areas. Finally, the realisation of instantaneous 3D scanning, could enable dynamic 3D motion scanning, whereby moving model are constructed from sequential scans. This could enabled advanced gait and movement analysis, enabling more sophisticated and accessible biomechanical assessment.

## 7 Conclusion

This study compared two 3D scanners that are commonly used in clinical practice and a novel prototype multi-camera photogrammetry-based 3D scanner. The results demonstrate that while the photogrammetry produced the scans with the highest accuracy and completeness, all three scanners provide a good level of accuracy and completeness that is sufficient for manufacture of custom foot orthoses when compared to the Artec Spider gold-standard control scanner. This study also demonstrates that the multicamera photogrammetry 3D scanner allows much faster 3D scan acquisition, which in addition to improving clinical workflows and efficiency, could enable better health care for people who cannot remain still for the duration required to use a current scanner.

## Acknowledgements

Not applicable.

## 10 Ethics approval and consent to participate

Ethics approval was obtained from the QUT University Human Research Ethics Committee (ethics approval number 6316). All participants provided signed consent for participation and image acquisition.

## 12 Authors’ contributions

JAT: Methodology, investigation, data curation, formal analysis, writing – original draft

AJT: Conceptualisation, methodology, data curation, formal analysis, software, Writing – review & editing

AJW: Supervision, methodology, project administration, writing – review & editing

RN: Methodology

ON: Resources, software

EP: Writing – review & editing

DH: Writing – review & editing

SKP: Conceptualisation, investigation, supervision, resources, methodology, writing – review & editing

MAW: Conceptualisation, supervision, methodology, writing – review & editing, funding support

## 13 Data Availability Statement

The data that support the findings of this study are available from the corresponding author, MAW, upon reasonable request and in alignment with the ethics approval from QUT.

## 14 Data Deposition

N/A

## 15 Biographical note

JAT: Joshua Taylor holds a Bachelor of Podiatry with First Class Honours and a Graduate Certificate in Podiatric Therapeutics, both from Queensland University of Technology (QUT). He works full-time in clinical practice on the Sunshine Coast, Queensland.

AJT: Alexander J Terrill is a lecturer, podiatrist, mechanical engineer and PhD candidate. His PhD research is at the interface of podiatry and engineering and applies advanced manufacturing technologies (3D scanning, computer aided design and engineering, and 3D printing) to improve the effectiveness, acceptability, and accessibility of personalised offloading treatment for people living with diabetes-related foot ulceration.

AW: Dr. Aaron Wholohan is a Lecturer in Podiatry at QUT. He holds both Bachelor’s and Master’s degrees in Podiatry, as well as a Ph.D. in foot rehabilitation. His research focuses on advancing orthotic manufacturing technologies and exploring foot biomechanics, particularly in relation to foot function and rehabilitation.

RN: Renee Nightingale holds a B.Eng (Hons) in Medical Engineering from QUT.

ON: Ollie Nagle is a mechatronics/aerospace engineer, with a diverse background in 3D CAD modelling, FDM/SLA 3D printing, robotics, computer vision and PCB design + fabrication.

EP: Dr Edmund Pickering is postdoctoral researcher in both the Biomechanics and Spine Research Group (BSRG) and the Biofabrication and Tissue Morphology (BTM) group at QUT. His research involves extensive image processing, mechanical testing and characterisation, 3D modelling and finite element simulation.

DH: Dr David Holmes is an Associate Professor in Mechanical Design & Manufacturing at QUT. Dr Holmes holds a B.Eng(Hons) in Mechanical Engineering and a Ph.D. in Computational Mechanics from James Cook University, with additional RPEQ and CPEng status in Mechanical Engineering Design. His research focusses on the development and application of advanced numerical methods as they apply to problems in flow, mechanical engineering design, and fatigue analysis.

SKP: Dr Sean Powell is a Senior Research Fellow in the BTM group and the School of Mechanical, Medical and Process Engineering at QUT. He holds a BAppSc(Physics) with first class Honors, and a Ph.D. also in physics. His research experience is in theoretical and computational modelling of particle dynamics and diffusion, and his current research is around 3D scanning, modelling and 3D printing for healthcare applications, including scanning hardware and software/AI for model processing and analysis.

MAW: Professor Maria A. Woodruff leads the BTM group at QUT and has 21 years’ experience in biomedical engineering since her undergraduate degree and PhD at the University of Nottingham, UK. Mia’s vision is of a future where the fabrication of patient-specific replacement tissue and devices are safe, cost-effective and routine and where 3D medical scanning modelling and printing can positively impact healthcare service delivery.

